# Role of TPM2 in Pediatric Restrictive Cardiomyopathy: Insights from Single-Nucleus RNA Sequencing and Proteomic Analysis

**DOI:** 10.1101/2025.08.01.25332843

**Authors:** Jie Liu, Zubo Wu, Cong Zhao, Qing Guo, Nianguo Dong, Peng Zhu, Jiawei Shi, Lin Wang, Hua Peng

**Affiliations:** Department of Pediatrics, Union Hospital, Tongji Medical College, Huazhong University of Science and Technology, Wuhan, China; Department of Cardiovascular Surgery, Union Hospital, Tongji Medical College, Huazhong University of Science and Technology, 1277 Jiefang Avenue, Wuhan, China

**Keywords:** RCM, Tropomyosin 2, SnRNA-seq, Children, Cardiomyocytes

## Abstract

Restrictive cardiomyopathy (RCM) is an uncommon pediatric condition characterised by diastolic dysfunction caused by myocardial stiffness, with preserved systolic function in the early stage. Its pathogenesis is linked to genetic mutations, metabolic defects, or fibrosis, but this process remains incompletely understood. Bioinformatics analysis indicated a crucial role of tropomyosin 2 (TPM2) in pediatric RCM. Using single-nucleus RNA sequencing (snRNA-seq) and proteomics, we identified molecular alterations in RCM hearts compared with controls, with a notable finding that TPM2 expression was markedly reduced in RCM patients. Functional assays showed that TPM2 knockdown in H9C2 cells promoted cell cycle progression (from G0/G1 to S phase), increased apoptosis, and enhanced cell migration. Subsequent western blot analysis confirmed alterations in cyclin-D1 and epithelial-mesenchymal transition (EMT) - related proteins following TPM2 silencing. These findings suggest that TPM2 may act as a cardioprotective factor and biomarker for pediatric RCM, thereby providing new therapeutic targets for this severe condition.

## Introduction

Restrictive cardiomyopathy (RCM) is characterized by impaired ventricular filling secondary to increased myocardial stiffness and pathologically elevated ventricular filling pressures, which results in restrictive cardiac physiology^1^. As the rarest form of cardiomyopathy, RCM poses particular diagnostic challenges for clinicians, who may be less familiar with its management compared to more prevalent cardiomyopathies. The condition typically manifests between the ages of 6 and 10 years; however, presentations have been documented across the lifespan, ranging from early infancy to adulthood, with no significant sex predilection. The broad age range of onset, combined with the rarity of the condition, often leads to diagnostic uncertainty-a phenomenon frequently observed in pediatric cases^2^.

RCM manifests in a wide range of clinical forms, from asymptomatic cases to overt heart failure (HF). The primary clinical manifestation of the condition is reduced exercise tolerance and fatigue, which result from decreased cardiac output. This is due to the left ventricle’s (LV) inability to adjust filling volumes during exertion, a condition that is further exacerbated by an elevated heart rate^3^. The symptoms of HF are non-specific and age-dependent. Infants typically present with feeding difficulties, tachypnea, sinus tachycardia, and diaphoresis. Children often have a history of recurrent lower respiratory tract infections, nocturnal wheezing, or a persistent cough. Adolescents may experience fatigue, dyspnea, or abdominal pain^4^.

Tropomyosin (TPM) is a sarcomeric actin-binding protein that critically regulates striated muscle contraction by modulating actin-myosin interactions. Beyond its canonical role in the sarcomere, TPM also governs cytoskeletal dynamics in migratory cells and serves as an essential regulator of myogenesis during early sarcomere assembly. The *Drosophila* genome encodes two TPM isofroms (Tm1 and Tm2), while humans express four (TPM1-4), each with distinct tissue-specific expression patterns: TPM1 predominates in cardiac muscle and type II fast-twitch skeletal fibers; TPM2 and TPM3 are enriched in type I slow-twitch muscle fibers.

Dominant pathogenic variants in TPM1, TPM2, and TPM3 are implicated in congenital myopathies^5-7^ and cardiomyopathies^8, 9^. Disease-associated mutations often disrupt TPM’s functional properties, including its stability, actin affinity, or calcium sensitivity, thereby driving the pathogenesis of these inherited disorders^10, 11^. Nevertheless, the precise spatiotemporal expression profile and functional contributions of TPM2 in cardiac development and disease remain incompletely characterized.

Single-cell RNA sequencing (scRNA-seq) has emerged as an indispensable tool in cardiovascular research—a field that demands deeper mechanistic insights, as heart disease remains the leading cause of global mortality with unresolved pathophysiological complexities. Given the large cellular volume of cardiomyocytes, nuclear transcriptomic profiling (single-nucleus RNA sequencing, snRNA-seq) offers superior cellular coverage and more representative characterization of cardiac tissue composition. By resolving the transcriptional profiles of individual cardiac cells, snRNA-seq has started to reveal previously inaccessible details about both physiological cardiac function and pathological remodeling, thereby creating new opportunities for mechanistic exploration and therapeutic development.

To systematically characterize the cellular landscape of RCM, we performed high-throughput snRNA-seq on 70,958 viable nuclei isolated from myocardial tissues of 3 pediatric RCM patients and 2 age-matched healthy controls. Using an integrative multi-modal bioinformatics approach, we identified cell type-specific transcriptional profiles, differentially expressed genes, and functionally enriched pathways distinguishing disease and control groups.

## Methods

### Sample

Diagnosis of RCM was established by echocardiography following the American Heart Association diagnostic criteria^12^. The etiology of pediatric RCM cases in this study remained undefined, consistent with previous classifications of idiopathic RCM. Patients were excluded from the study if they received a confirmed diagnosis of cardiomyocyte storage diseases or inherited metabolic disorders. Clinical data for all participants were extracted from electronic hospital records. Donors underwent comprehensive clinical evaluations and medical history reviews, which revealed no evidence of structural heart disease.

Left ventricular (LV) myocardial tissue specimens from RCM patients were collected during heart transplantation (HT), immediately flash-frozen in liquid nitrogen, and stored at −80°C. Control samples consisted of flash-frozen donor heart tissues obtained from pre-transplantation myocardial biopsy specimens.

The utilization of human tissues for research purposes was approved by the Institutional Review Boards (IRBs) of both the Gift-of-Life Donor Program and Wuhan Union Hospital, in accordance with the ethical principles established by the Declaration of Helsinki. This study received formal approval from the Ethics Committee of Union Hospital [ethical approvals: KY2017-323]. Written informed consent was obtained from all participants, explicitly detailing the research objectives of tissue donation.

### SnRNA-seq analysis

Human myocardial specimens were obtained from 3 RCM patients undergoing cardiac transplantation and 2 age-matched healthy donors undergoing diagnostic myocardial biopsy (Table S1-S2). Single nuclei were isolated from cardiac tissues and processed for snRNA-seq utilizing the 10×Genomics Chromium platform. Subsequent bioinformatics analysis enabled identification of distinct cellular subpopulations and their characteristic differentially expressed gene profiles, which served as reference data^13^.

Top variable genes were identified, with highly variable genes selected using Scanpy’s **pp.highly_variable_genes** function. Dimensionality reduction was performed via principal component analysis (PCA; **tl.pca** Scanpy). Cell clustering was conducted using graph-based methods (**pp.neighbors** in Scanpy), and results were visualized in 2D/3D using Uniform Manifold Approximation and Projection (UMAP; **tl.umap**). Cluster-specific marker genes were determined using Seurat’s **FindAllMarkers** function (Wilcoxon rank-sum test, adjusted *P* < 0.05, |log∼2∼FC| > 0.25). Enrichment analysis of marker genes was conducted using ClusterProfiler, assessing overrepresentation in Gene Ontology (GO) terms and KEGG pathways via hypergeometric testing.

### RNA-sequencing and transcriptional analyse

Human cardiac tissue specimens were obtained from seven pediatric RCM patients during indicated heart transplantation procedures and three age-matched healthy donors during diagnostic myocardial biopsies (Supplemental Table 1-2). Subsequent RNA and protein isolation was performed as described below.

### RNA Isolation, Quality Assessment, and Sequencing

Total RNA was extracted from cardiac tissue using TRIzol® Reagent (Invitrogen, Carlsbad, CA, USA). RNA purity and concentration were assessed using a NanoDrop™ spectrophotometer (Thermo Fisher Scientific, Waltham, MA, USA) and quantified via Qubit® 4.0 Fluorometer (Thermo Fisher Scientific). RNA integrity and quantity were further evaluated using an Agilent 2100/4200 Bioanalyzer system (Agilent Technologies, Santa Clara, CA, USA).

Following stringent quality control, RNA libraries were constructed for each sample. Purification and fragment size selection were performed using Hieff NGS® DNA Selection Beads (Yeasen Biotechnology, Shanghai, China). Libraries were then amplified and enriched via polymerase chain reaction (PCR). Finally, high-throughput sequencing was carried out on a DNBSEQ-T7 platform (MGI Tech, Shenzhen, China) employing combinatorial probe-anchor synthesis (cPAS) technology.

### Protein extraction and tag label-free mass spectrometry analysis

Cardiac tissue samples were homogenized in lysis buffer containing 2.5% SDS and 100 mM Tris-HCl (pH 8.0). Following centrifugation, proteins in the supernatant were precipitated by adding four volumes of pre-cooled acetone. Subsequent steps included protein pellet dissolution, reduction, and alkylation. Protein concentration was determined using the Bradford assay, and the resulting peptide eluate was lyophilized under vacuum and stored at -20℃ for further analysis. Liquid chromatography-tandem mass spectrometry (LC-MS/MS) was performed using a Q Exactive Plus mass spectrometer (Thermo Scientific) coupled with an Easy-nLC 1200 system.Raw LC-MS/MS data were processed using MaxQuant (version 1.6.6) with the Andromeda search algorithm. Spectra were matched against the UniProt Human proteome database for protein identification and quantification.

### Cell culture

H9C2 rat cardiomyoblasts (Warner Bio Co., Ltd.) were maintained in Dulbecco’s Modified Eagle Medium (DMEM; Gibco, Leicestershire, UK) supplemented with 10% fetal bovine serum (FBS; Gibco) and 1% penicillin-streptomycin (Gibco) at 37°C in a humidified 5% CO₂ incubator.

### Cell transfection

H9C2 cell were resuspended in complete culture medium and plated at a density of 5×10^5^ cells/well in 6-well plates. After overnight culture, the medium was replaced with serum-free DMEM. Cells were then divided into four experimental groups: siRNA negative control (NC); si-TPM2-001; si-TPM2-002 and si-TPM2-003. All siRNA constructs were designed and synthesized by *Tsingke Biotech* (Beijing). The sequences of si-RNA were as follows: NC, sense: 5’-UUCUCCGAA CGUGUCACGU(dT)(dT)-3’, antisense: 5’-ACGUGACACGUUC GGAG AA(dT)(dT)-3’; si-TPM2-001, sense: 5’-GGAGAUGCAGCUGAAAGAA(dT) (dT)-3’, antisense: 5’-UUCUUUCAGCUGCAUCUCC(dT)(dT)-3’; si-TPM2-002, sense: 5’-GACAAAUACGAAGAAGAGA(dT)(dT)-3’, antisense: 5’-UCUCUUCUU CGUAUUUGUC(dT)(dT) -3’; si-TPM2-003, sense: 5’-CGAAGAAGAGAUCAAAC UU(dT)(dT)-3’, antisense: 5’-AAGUUUGAUCUCUUCUUCG(dT)(dT)-3’. Cell transfection was performed as previously described^14^. Briefly, H9C2 cells were seeded at a density of 5×10^5^ cells per well into 6-well plates and cultured overnight. Subsequently, the cell medium was changed to serum-free medium, and the cells were subjected to transfection with 50nM NC or 50nM si-TPM2 (001/002/003) at 24 ± 2℃ for 15 min using Lipofectamine® 3000 (Thermo Fisher Scientific, Inc.) in accordance with the manufacturer’s protocol. Cells in the blank control group were cultured in medium. Following transfection for 6 h, the medium was replaced with complete culture medium. After culture for another 48 h, the transfection efficiency was evaluated using RT-qPCR and WB.

### Cell apoptosis assay

The transfected H9C2 cells were harvested at 48 hours post-transfection via enzymatic dissociation with 1×trypsin-EDTA solution (Invitrogen). Subsequently, the cell suspension was adjusted to a density of 1×10^6^ cells/mL in Annexin V binding buffer and dual-stained with fluorescein isothiocyanate (FITC)-conjugated Annexin V and propidium iodide (PI) utilizing the FITC Annexin V Apoptosis Detection Kit I (BD Biosciences, Franklin Lakes, NJ). Following a 15-minute incubation period at ambient temperature under light-protected conditions, cellular fluorescence was quantified by flow cytometric analysis performed on a FACScan instrument (BD Biosciences) with accompanying Cell Quest Pro software (BD Biosciences) for data acquisition and analysis.

### TUNEL staining

Cell climbing slides were fixed and permeabilized, then incubated with TdT reaction buffer (MK1018, Boster) at 37°C for 2h in a humidified chamber. After PBS washing, nuclear staining was performed with 4’,6-diamidino-2-phenylindole (DAPI) prior to fluorescence microscopic observation.

### Cell migration assay

H9C2 cell migratory capacity was assessed using a Transwell migration assay with polycarbonate membrane inserts (8 μm pore size; Guangzhou Jet Bio-Filtration Co., Ltd). Transfected cells (1 × 10^5^cells/well) were suspended in complete medium and seeded into the upper chamber, while the lower chamber was filled with medium containing 20% fetal bovine serum (FBS) as a chemoattractant. After 48 h of incubation at 37°C under 5% CO2, migratory cells adhering to the lower membrane surface were fixed with 4% paraformaldehyde (Sinopharm Chemical Reagent Co., Ltd.) for 20 min at room temperature. Subsequently, cells were stained with 0.5% crystal violet (Beyotime Institute of Biotechnology) for 10 min at ambient temperature. Migrated cells were then quantified and imaged under an inverted light microscope (Olympus Corporation).

### Cell cycle

Transfected H9C2 cells (1×10⁶) were harvested and washed twice with ice-cold phosphate-buffered saline (PBS). Cell fixation was performed using 70% ethanol at -20°C for 2 hours. After two additional PBS washes, cells were resuspended in 1 mL PBS containing 50 μg/mL RNase A (Sigma-Aldrich) and incubated at 37°C for 1 hour to ensure complete RNA digestion. Following another wash step, cells were stained with 50 μg/mL PI for 10 minutes at room temperature in the dark. Cell cycle distribution was analyzed by flow cytometry (FACS Calibur, BD Biosciences) based on differential DNA content staining patterns.

### RT-qPCR

Total RNA was isolated from lung tissues using TRIzol reagent (Sigma-Aldrich) according to the manufacturer’s protocol. RNA purity and concentration were verified by spectrophotometry (A260/A280 ratio >1.8). First-strand cDNA was synthesized from 1 μg total RNA using the ALL-in-One cDNA Synthesis Kit (GeneCopoeia). Quantitative real-time PCR (RT-qPCR) was performed on a Bio-Rad CFX96 system in 10μl reactions containing: 1μl cDNA template, 5μl 2× ALL-in-One qPCR Mix (GeneCopoeia), 0.2μl each of forward and reverse primers (human TPM2 forward: 5’-GAGAGGTCTGTGGCAAAGTTGG-3’, reverse: 5’-GGAGGTGATGTCATTGAGTGCG-3’; human GAPDH forward: 5’-T GACTTCAACAGCGACACCCA-3’, reverse: 5’-CACCCTGTTGCTGTAGCCAAA -3’; rat TPM2 forward: 5’-AAGGGGACAGAGGATGAG-3’, reverse: 5’-CTTTCTCA GCCTCCTCCA-3’; rat GAPDH: forward: 5’-TT GGCCGTATTGGCCGC-3’, reverse: 5’-GTGCCATTGAACTTGCCGTG -3’), and 3.6μl nuclease-free water. The thermal cycling protocol consisted of: initial denaturation at 95℃ for 10 min; 39 cycles of 95℃ for 10 sec, 55℃ for 30 sec, and 72℃ for 30 sec; final extension at 65℃ for 5 sec; followed by melt curve analysis (95℃ for 5 sec, 65℃ to 95℃ in 0.5℃ increments). Relative gene expression was calculated using the 2-ΔΔCt method with GAPDH as the endogenous control.

### Western blotting

Lung tissues were homogenized in RIPA lysis buffer (Thermo Fisher Scientific) supplemented with protease and phosphatase inhibitors (Complete Protease Inhibitor Cocktail, Sigma-Aldrich). Protein concentrations were determined using a BCA assay kit (Invitrogen) following the manufacturer’s protocol. Protein samples were denatured by boiling at 95°C for 10 min in Laemmli buffer. Equal amounts of protein (20μg per lane) were separated by SDS-PAGE (10% gel) and electrophoretically transferred to nitrocellulose membranes (0.45μm pore size). Membranes were blocked with 3% bovine serum albumin (BSA) in TBST (Tris-buffered saline with 0.1% Tween-20) for 1 h at room temperature, followed by overnight incubation at 4℃ with the following primary antibodies: rabbit anti-TPM2 (1:1000; 11038-1-AP; Proteintech), mouse anti-Cyclin-D1 (1:10000; 60186-1-Ig; Proteintech), rabbit anti-Vimentin (1:1000; 5741; Cell Signaling Technology); rabbit anti-beta-catenin (1:1000; 8480; Cell Signaling Technology); rabbit anti-TGF-beta1 (1:10000; 81746-2-RR; Proteintech); Rabbit anti-alpha SMA (1:10000; ab124964; abcam); rabbit anti-Snail (1:1000; 3879; Cell Signaling Technology); mouse anti-Alpha-Tubulin (1:200000; 66031-1-Ig; Proteintech) and mouse anti-GAPDH (1:50,000; 60004-1-Ig, Proteintech). After washing with TBST, immunoreactive bands were detected using an Odyssey CLx infrared imaging system (LI-COR Biosciences) and quantified using ImageJ software.

### Immunohistochemistry

For immunohistochemistry, paraffin sections were deparaffinized twice in 100% xylene for 20 min and rehydrated by incubation in decreasing concentrations of ethanol (100%, 95%, 85%, and 60% for 5 min each). Antigen retrieval was then performed using Tris-EDTA (95℃ for 20 min), followed by incubation in 0.3% (vol/vol) H2O2 in Tris-buffered saline (TBS) for 30 min at room temperature. The sections were permeabilized in 0.5% (vol/vol) Triton X-100 (in TBS) for 30 min and then blocked in 3% bovine serum albumin (in TBS) for 1 h at room temperature, followed by incubation with primary antibodies at 4℃ overnight. The following primary antibodies were used for immunohistochemistry: rabbit anti-TPM2 (1:200; 11038-1-AP; Proteintech). Sections were then washed three times in TBS for 5 min before incubation with goat anti-rabbit/mouse horseradish peroxidase (HRP)-labeled polymer (KIHC-5, Proteintech, Wuhan, China) for 1 h at room temperature. The slices were washed three times in TBS for 5 min and then stained in 3,3-diaminobenzidine (DAB) staining solution (KIHC-5, Proteintech, Wuhan, China) for 10 min and then quickly washed three times in PBS for 5 min, followed by counterstaining with a hematoxylin kit (PMK-341, Bioprimacy, Wuhan, China). The stained slices were dehydrated in increasing concentrations of ethanol 60%, 85%, 95%, and 100% for 5 min each, followed by another wash in 100% ethanol for 5 min. The slides were then immersed in 100% xylene for 20 min twice. Finally, the slides were mounted with neutral balsam. Immunohistochemistry images were visualized using a light microscope (Olympus Corporation).

### Sirius red staining

Paraffin-embedded tissue sections were subjected to standard deparaffinization through xylene immersion and graded ethanol hydration. Collagen fibers were specifically stained using 0.1% saturated picric acid-Sirius Red solution (Sigma-Aldrich) for 60 minutes at room temperature, followed by thorough washing under running distilled water for 5 minutes. Nuclear counterstaining was performed using Harris hematoxylin. Sections were then dehydrated through an ascending ethanol series, cleared in xylene, and permanently mounted with synthetic resin mounting medium. Histological evaluation was performed using both bright-field and polarized light microscopy (Olympus BX53) to assess collagen fiber distribution and birefringence patterns.

### Statistical analysis

Continuous variables are presented as mean ± standard deviation (SD). Statistical analyses were performed using GraphPad Prism version 9.3.0 (GraphPad Software, San Diego, CA). Between-group comparisons were analyzed by two-tailed unpaired Student’s t-test. A P-value <0.05 was considered statistically significant.

## Results

### 1. Proteomic changes of RCM

A quantitative proteomic profiling assay was performed using tandem mass tags (TMT) on human heart lysates, which were derived from seven pediatric patients with RCM and three donor myocardial biopsies. Tropomyosin 2 (TPM2), a member of the tropomyosin-actin filament binding protein family, showed a significant decrease in pediatric patients with restrictive cardiomyopathy (RCM) compared with the control group (Figure 1A). Gene Ontology (GO) analysis for biological processes (BP) indicated that cytoplasmic translation, cellular respiration, mitochondrial respiratory chain complex assembly, regulation of focal adhesion assembly and regulation of cell-substrate junction organization and assembly were all significantly enriched. GO analysis for cellular components (CC) revealed that focal adhesions and cell-substrate junctions were among the differentially expressed cellular components (Figure 1B). It was observed that TPM2 forms a dense interaction cluster with cardiac sarcomeric proteins (TNNI3 and TNNI2, with variants identified in children with sporadic RCM), suggesting its pivotal role in pediatric RCM (Figure 1C).

**Figure 1.**
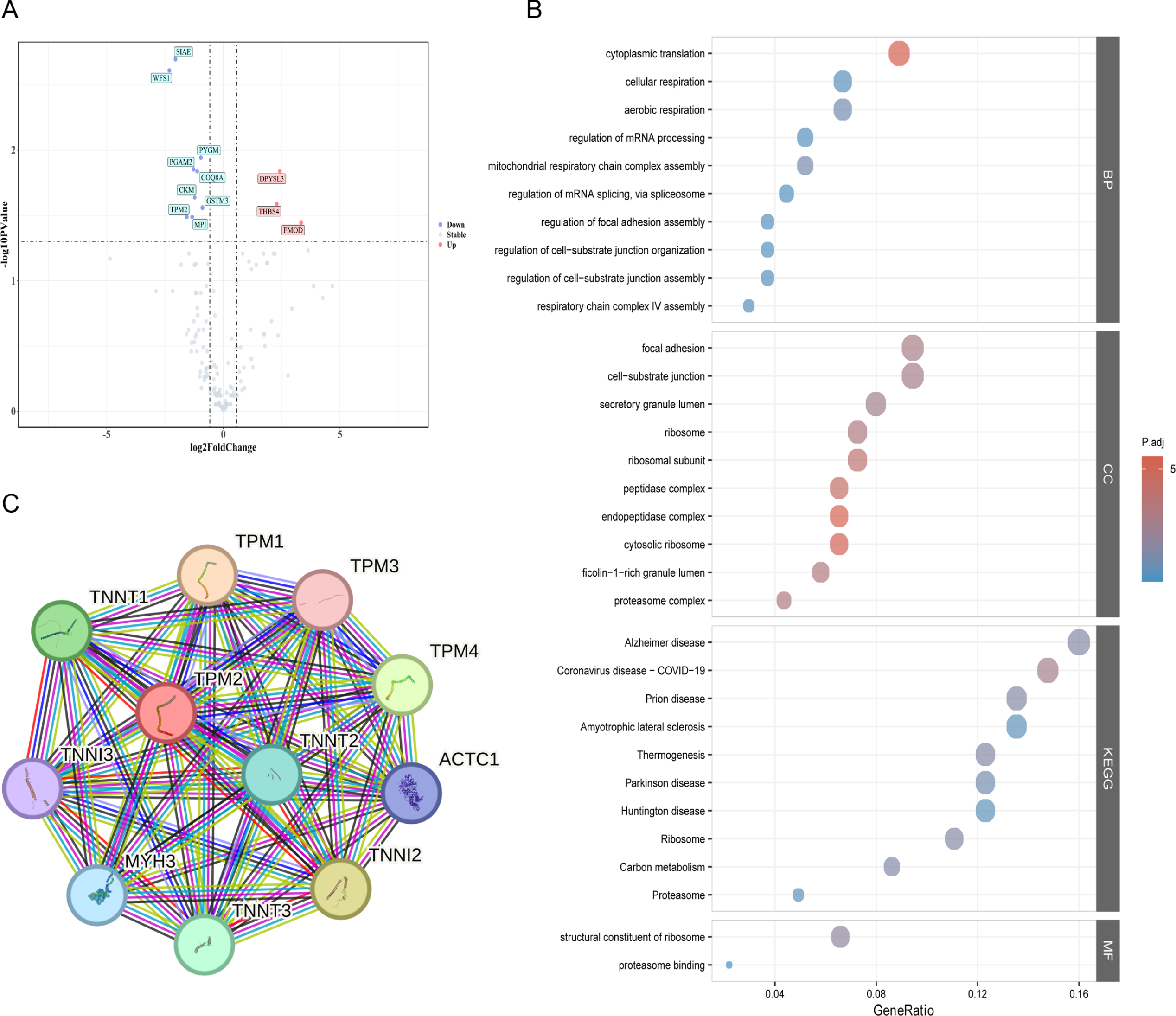
Characterization of comparative proteome analysis in pediatric RCM and control hearts. A. Volcano plot displaying differentially expressed proteins in left ventricular myocardial tissues from pediatric RCM hearts compared with control hearts. Each dot represents a protein; proteins with significantly upregulated expression are labeled in red, and those with significantly downregulated expression are labeled in blue (fold change≥ 1.2, P < 0.05). B. GO (Gene Ontology) and KEGG (Kyoto Encyclopedia of Genes and Genomes) pathway enrichment analyses of differentially expressed proteins. The top 10 enriched GO and signaling pathways (KEGG) are shown, with the size of each bubble representing the number of proteins enriched in the corresponding term/pathway and the color indicating the significance of enrichment (P value). C. Expression profiles of differentially expressed proteins related to TPM1, TNNI3 and TNNI2.

### 2. Optimization and Recharacterization of Single-Cell Transcriptomic Alterations in RCM

Single nucleus RNA-sequencing (snRNA-seq) was performed in triplicate on left ventricular (LV) samples from four individuals, including two children with RCM and two subjects with non-cardiac disease. All RCM patients were diagnosed with advanced cardiomyopathy, which required heart transplantation. After removing low-quality nuclei, 25,742 nuclei were retained and further aggregated into nine clusters based on transcriptional similarity (Figure 2A-B). The dominant cell types identified included cardiomyocytes, fibroblasts, endothelial cells, myeloid cells, pericytes, and T cells. Compared with normal heart, RCM hearts showed a statistically significant reduction in cardiomyocytes, along with an increase in fibroblasts and endothelial cells. Further analysis of the cardiomyocyte cell cycle revealed that the proportion of cells in the G1 phase significantly decreased in RCM group, while the proportion of cells entering the S phase increased, indicating a potential cell cycle arrest (Figure 2C-D).

**Figure 2.**
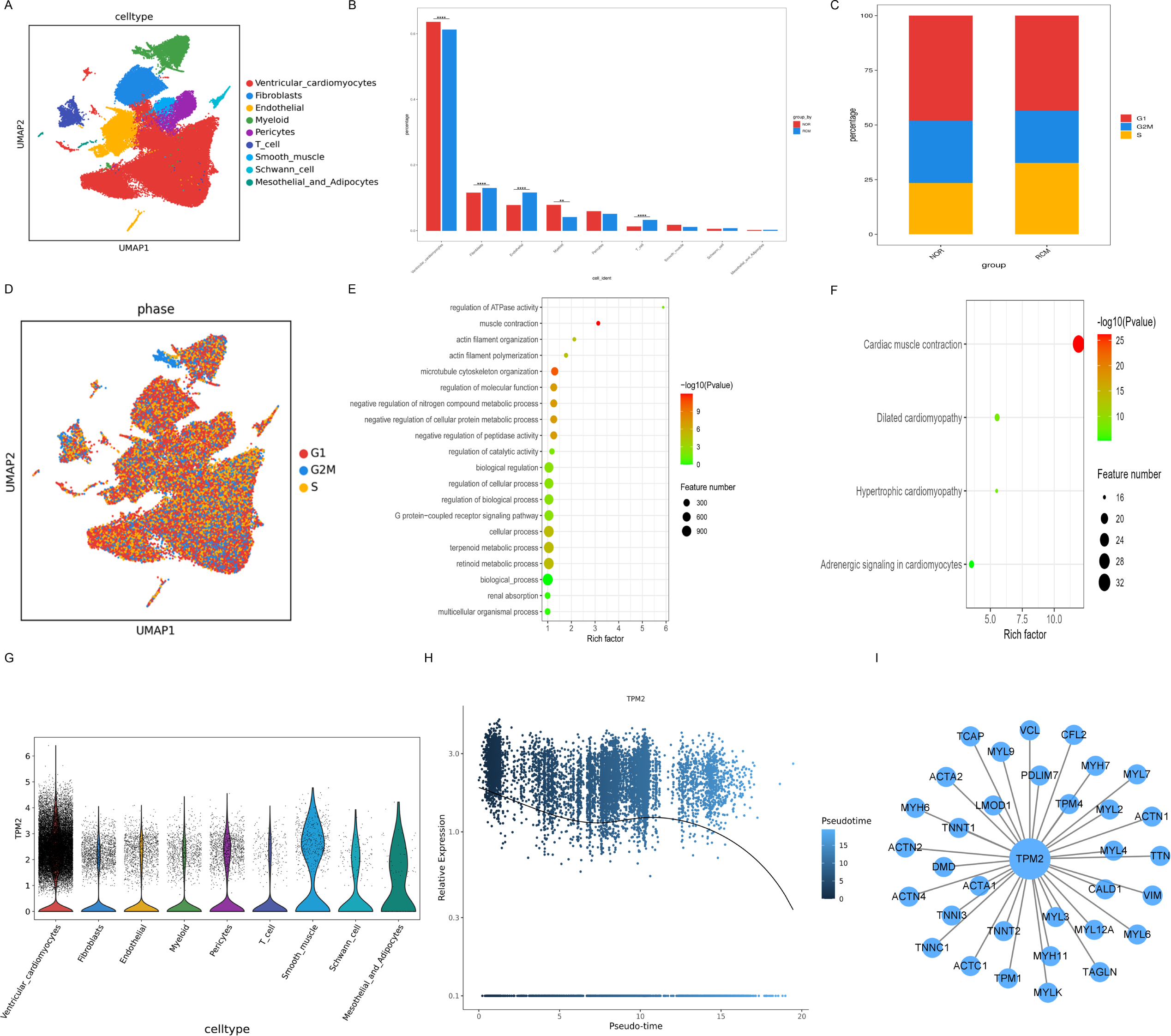
Single-cell sequencing analysis of cardiac tissues in pediatric RCM and control hearts. **A**. Uniform manifold approximation and projection (UMAP) plot of the multi-omics dataset. Cell types are annotated based on marker genes. **B**. Comparison of relative proportions of cell types between RCM and control groups. The bar chart shows the percentage of each cell type in the total cell population of each group. **C**. Bar plot displaying the cell cycle phase distribution of cardiomyocytes across all samples. The y-axis represents the proportion of cardiomyocytes in G1, G2M, and S phases. **D**. UMAP plot of multi-omics dataset, with cells colored according to their predicted cell cycle classification (G1, G2M, and S phase). **E, F**. Bubble chart showing GO (**E**) and KEGG (**F**) enrichment terms of differentially expressed genes in cardiomyocytes cluster. Bubble size indicates the number of genes enriched in the corresponding term, and color represents the enrichment significance (adjusted P value). **G**. Violin plots showing the expression level of TPM2 in different cell types. The x-axis represents cell origins, and the y-axis represents normalized expression values of TPM2. **H**. Time-series analysis of TPM2 expression in cardiomyocytes. The line graph shows the dynamic changes in TPM2 expression during cardiomyocyte development or aging. **I**. Protein-protein interaction network diagram. Nodes represent proteins, and edges indicate interactions between proteins; the network is centered on TPM2 and its interacting partners.

A study was conducted to screen genes specifically expressed in cardiomyocyte clusters. These genes were then used to assess enrichment in biological processes through gene ontology analysis for the corresponding gene sets (Figure 2E). To identify systematic patterns of gene dysregulation, pathway enrichment analyses were performed based on differential gene expression between RCM and control hearts (Figure 2F). Notably, we observed significant dyregulation of several pathways in RCM cardiomyocytes, including the regulation of ATP activity, muscle contraction, actin filament organization, actin filament polymerization and microtubule cytoskeleton organization. The specific gene marker set for cardiomyocytes was enriched in pathways related to cardiac muscle contraction and adrenergic signaling in cardiomyocytes, suggesting cardiac damaged during RCM progression.

To clarify the precise role of TPM2 in RCM progression, we characterised the spatial organisation of TPM2 over time. This study revealed that TPM2 is predominantly expressed in cardiomyocytes and smooth muscle cells (Figure 2G). The proposed time-series analysis showed that TPM2 expression was most abundant in naive cardiomyocytes, remained stable at the mature stage, and decreased with aging (Figure 2H). The protein-protein interaction (PPI) plot demonstrated a close association between TPM2 and TNNI3 genes (Figure 2I). It has been established that mutations in the TNNI3 gene are a significant genetic factor contributing to RCM in the pediatric population.

### 3. TPM2 expression in RCM

As indicated by the results of the proteome analysis, TPM2 protein expression was decreased. To validate these observations, immunoblot analysis was performed on left ventricular myocardial proteins from children with RCM and the control group. Compared with the control group, TPM2 protein expression in the RCM group was significantly downregulated, while no obvious changes in TPM2 mRNA expression were observed (Figure 3A-C). Immunohistochemical analysis of ventricular myocardium showed that TPM2 was predominantly expressed in ventricular muscle cells and vascular endothelial cells, and its protein expression in the RCM group was lower than that in the control group (Figure 3D). Histological examination of Sirius Red-stained samples from the RCM and control groups revealed disorganized arrangement of cardiac cells and collagen fiber deposition in RCM (Figure 3E). Polarization microscopy of LV myocardial tissue revealed that type III collagen fibers were predominant in control myocardial tissue sections. These green fine fibers exhibited weak birefringence, while type I collagen fibers were less prevalent, appearing as red or yellow coarse fibers. Type I collagen fibers coexisted with type III collagen fibers, and the latter showed strong birefringence. In the myocardium of the RCM group, type I collagen fibers were significantly increased, characterized by a coarse red or yellow appearance, strong refractive index, and obviously disordered arrangement (Figure 3F).

**Figure 3.**
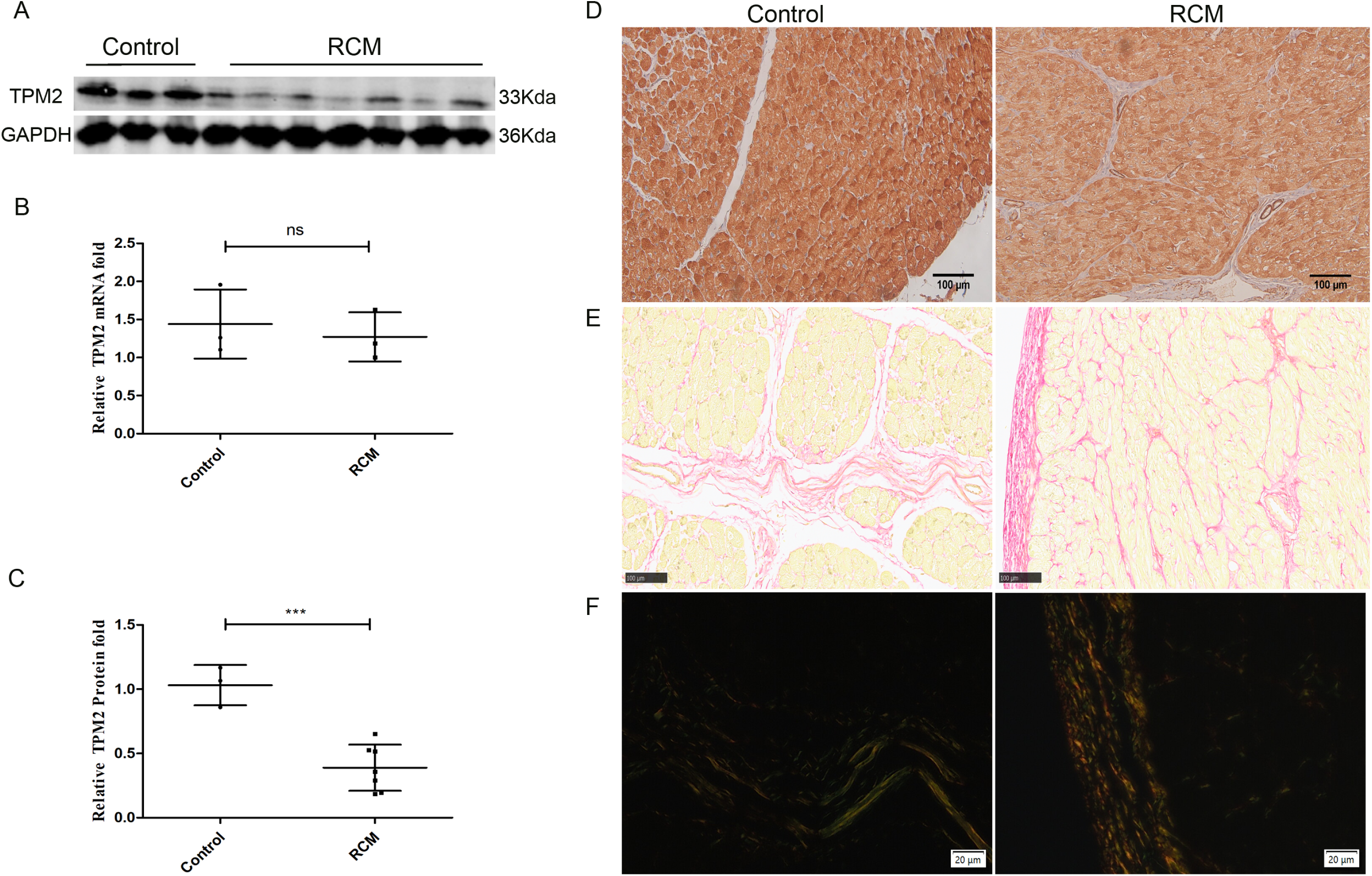
TPM2 mRNA and protein expression in pediatric RCM and control hearts. **A**. Representative blots showing TPM2 protein expression in RCM and control hearts. **B**. TPM2 mRNA expression in RCM and control hearts. The histogram displays the relative mRNA expression level of TPM2. **C**. Semi-quantification of TPM2 protein expression. The histogram shows the relative protein expression level of TPM2 normalized to the internal reference. **D**. Left ventricular myocardial tissue sections from RCM and control hearts stained with anti-TPM2 antibody. Positive staining is indicated by brown color. **E, F**: The content and distribution of collagen in human left ventricular tissues observed by conventional light and polarization microscopy combined with image analysis after picrosirius red staining. Red or yellow signals indicated coolagen fibers. Data are presented as mean ± SEM. ***P < 0.001 vs. control; ns, not significant. Scale bar: 100 μm (D and E); 20 μm (F).

### 4. Effects of TPM2 on cell apoptosis

To investigate the functional role of TPM2 in RCM, we utilized H9C2 cells. Three individual TPM2-targeting small interfering RNAs (siRNAs) were designed and synthesized. After screening, transfection efficiency was validated at both the mRNA and protein levels (Figure S1). Three independent TPM2-knockout siRNAs (siRNA-TPM2-001, si-TPM2-002 and si-TPM2-003) and a negative control (NC) were used for functional validation of TPM2 in H9C2 cells. The effects of TPM2 on H9C2 cell apoptosis was evaluated using flow cytometry (Figure 4A) and TUNEL staining (Figure 4B). Flow cytometry results showed that the apoptosis rates in the control, si-TPM2-001, si-TPM2-002 and si-TPM2-003 groups were 11.07 ± 0.7985%, 20.62 ± 2.858%, 17.18 ± 0.7879% and 14.67 ± 0.2713%, respectively. These results demonstrated that the cell apoptosis rate in the TPM2-silenced groups was significantly higher than that in the control group (Figure 4C-D). TUNEL staining was further performed to confirm the findings from flow cytometry. Collectively, these results indicated that TPM2 knockdown promoted H9C2 cell apoptosis compared with control cells.

**Figure 4.**
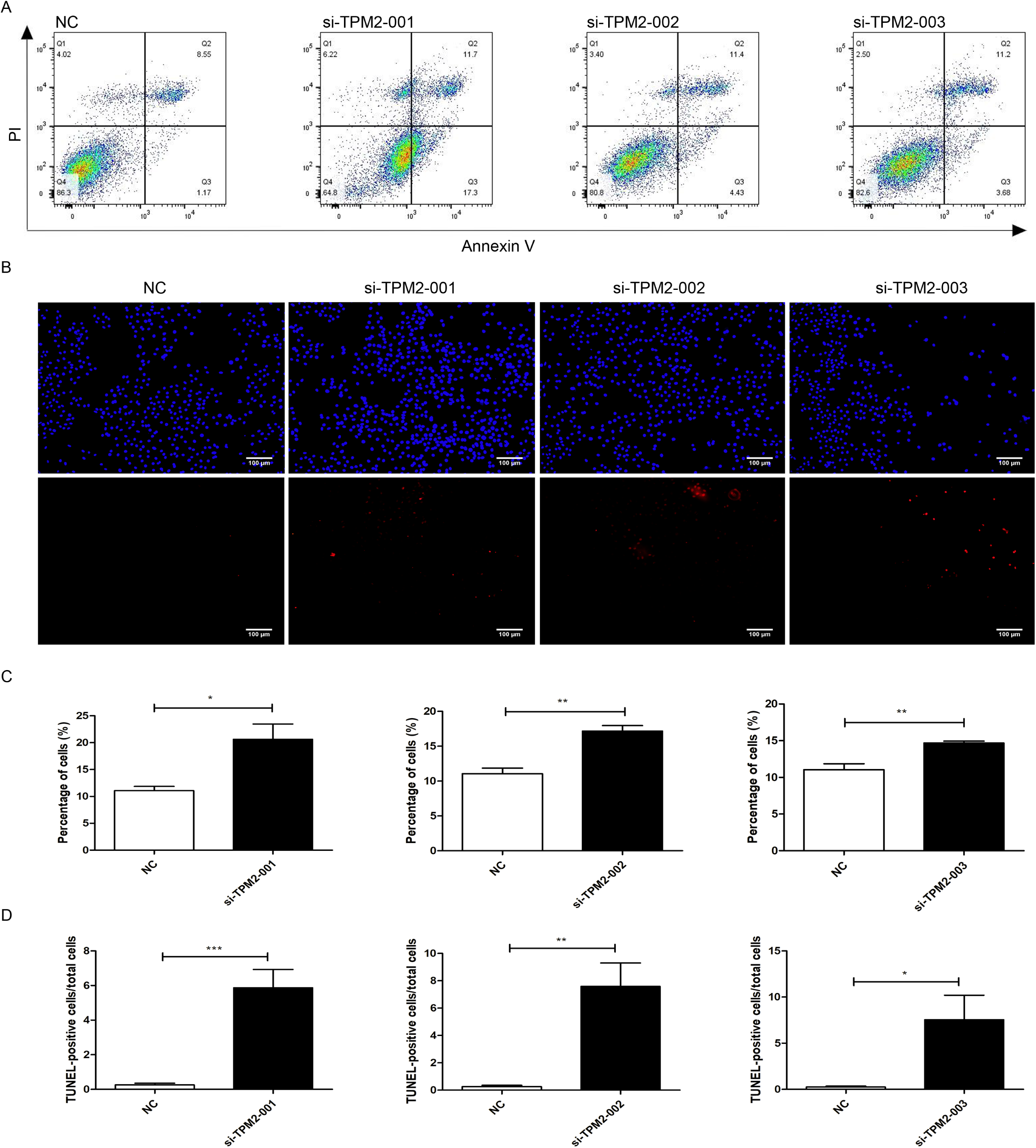
Effects of TPM2 on H9C2 cell apoptosis. H9C2 cell apoptosis was evaluated via (**A**) flow cytometry, with results (**C**) quantified. (**B**) TUNEL staining of H9C2 cells and (**D**) quantitative analysis of each treatment group. Blue fluorescence represents 4′6-diamidino-2-phenylindole-stained nuclei and red fluorescence indicates TUNEL - positive cells. Data are presented as mean ± SEM (n = 4 independent experiments). *P <0.05 vs. NC; **P<0.01 vs. NC; ***P<0.001 vs. NC; si-TPM2, small interfering RNA targeting TPM2; NC, negative control. Scale bar: 100 μm.

### 5. Effects of TPM2 on cell cycle

Flow cytometry was performed to analyze the cell cycle distribution of the si-TPM2 and control groups. As shown in the figure, the number of cells in G0G1 phase was significantly decreased in the si-TPM2 treatment group compared with the control group, while the number of cells in the S phases were significantly increased. These results indicate that TPM2 knockdown can induce H9C2 cells to transition from the G0G1 phase to the S phase (Figure 5A-B). Subsequently, we investigated the role of TPM2 in regulating cyclin-D1 protein expression. The cyclin-D1 protein level was found to be decreased in H9C2 cells transfected with si-TPM2 compared with the control group (Figure 5C-D).

**Figure 5.**
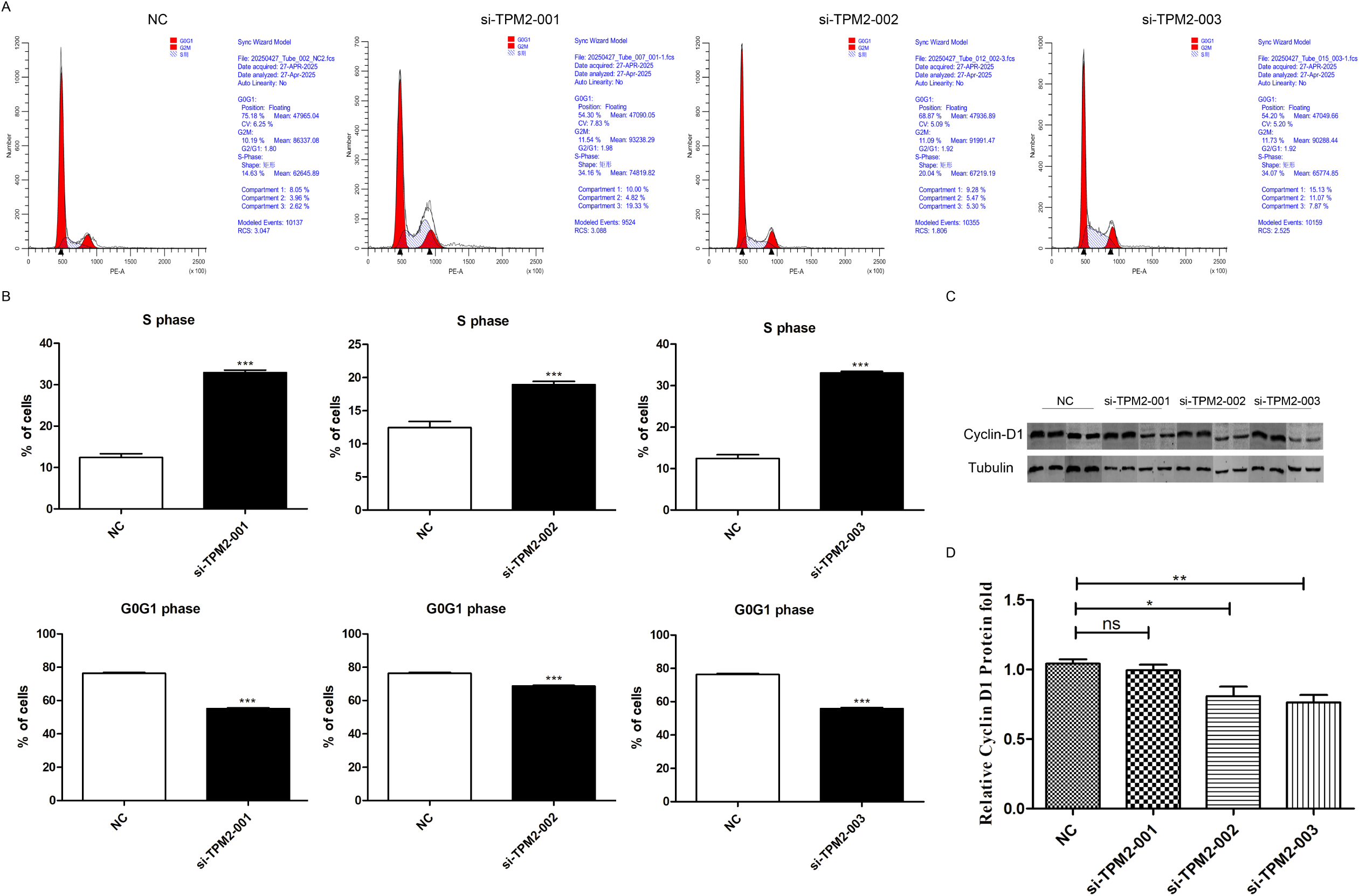
Effects of TPM2 on H9C2 cell cycle. H9C2 cells were labeled with propidium iodide and analyzed by flow cytometry (FACS). (A) A representative FACS histogram. (B) Quantification of cell frequency in G0G1 and S phase. (C) Representative blots and (**D**) quantification of Cyclin-D1 protein expression in the si-TPM2 and NC groups. Data are presented as mean ± SEM (n = 5 independent experiments). *P< 0.05 vs. NC; **P < 0.01 vs. NC; ***P < 0.001 vs. NC; si-TPM2, small interfering RNA targeting TPM2; NC, negative control.

### 6. Effects of TPM2 on cell migration

A Transwell assay was performed to evaluate the effect of TPM2 on cell migration (Figure 6A). Compared with the control group, the number of migrated cells in the si-TPM2 groups was significantly increased (Figure 6B), suggesting that TPM2 knockdown promoted migration in H9C2 cells. Western blotting (WB) was conducted to explore the molecular mechanisms underlying the effect of TPM2 knockdown on H9C2 cell migration. Compared with the control group, the protein expression levels of TGF-β1, α-SMA, and vimentin were significantly increased in the si-TPM2 groups. Among the epithelial-mesenchymal transition-related protein, Snail expression was elevated only in the si-TPM2-001 group. In contrast, the protein expression level of β-catenin was decreased (Figure 6C-H).

**Figure 6.**
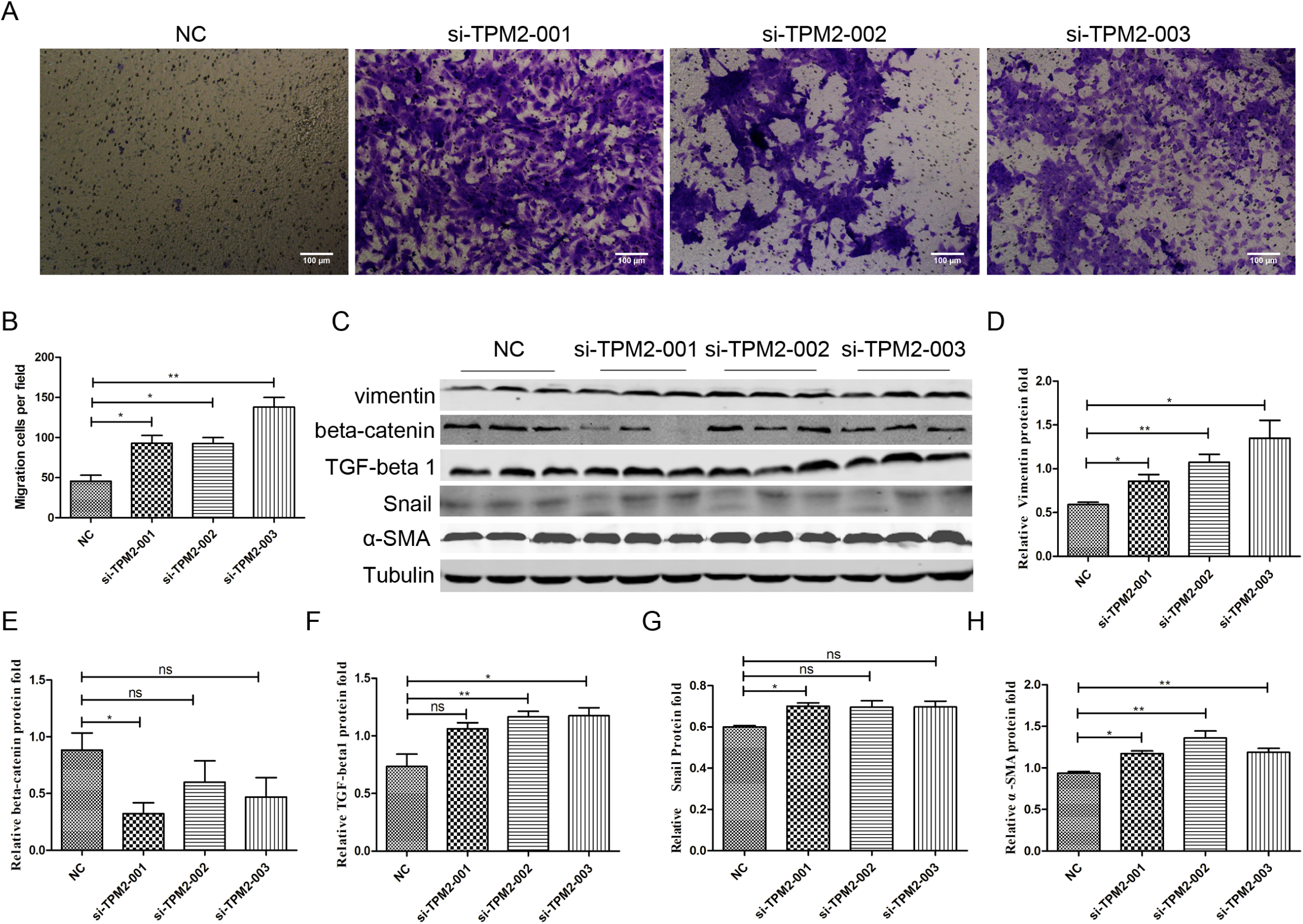
Effects of TPM2 on H9C2 cell migration. (**A**) H9C2 cell migration was assessed using Transwell assays and (**B**) quantified. (C) Representative blots and (D) quantification of TGF-β1, α-SMA and epithelial-mesenchymal transition (EMT) - related protein expression in the si-TPM2 and NC groups. Data are presented as mean ± SEM. *P<0.05 vs. NC; **P<0.01 vs. NC; ns, negative significant; si-TPM2, small interfering RNA targeting TPM2; NC, negative control. Scale bar: 100 μm.

## Discussion

Restrictive cardiomyopathy (RCM) accounts for 3%-5% of cardiomyopathy cases in pediatric populations, characterized by diastolic dysfunction, impaired ventricular filling, and atrial enlargement. Gaining insights into the molecular mechanisms underlying pediatric RCM and identifying novel therapeutic targets are therefore of critical importance. In this study, we investigated cellular changes and characteristics of relevant cell subpopulations during RCM progression in pediatric patients by integrating proteomic sequncing and single-nucleus RNA sequencing (snRNA-seq) data. Our findings revealed compositional shifts in cell types and subtypes between RCM-affected and control hearts, with a notable reduction in ventricular cardiomyocytes and an increase in fibroblasts in RCM hearts. These observations suggest that either cardiomyocytes or cardiac fibroblasts—key contributors to cardiac diastolic dysfunction—may act as a crucial regulators of RCM progression.

Proteomic analysis indicated that tropomyosin 2 (TPM2) may play an important role in RCM. Tropomyosins (TPMs) are a family of actin-binding proteins with highly tissue-specific expression patterns. Their primary function is to stabilize and integrate actin filaments, and they are also involved in cell migration and morphological changes^15, 16^. Previous studies using knockout mouse models have shown that TPM2 is essential for development and survival, as homozygous TPM2 deletion results in embryonic lethality. While TPM2 is predominantly expressed in skeletal muscle, its role in cardiac tissue remains poorly understood. In the current study, proteomic analysis identified downregulated TPM2 expression in pediatric RCM hearts, a finding confirmed by Western blotting. We therefore hypothesized that aberrant TPM2 expression may be associated with pediatric RCM. SnRNA-seq results further showed that TPM2 is mainly expressed in ventricular myocytes and smooth muscle cells within left ventricular myocardial tissues of pediatric RCM patients. We thus used H9C2 cells to explore TPM2 function. Aberrant TPM2 expression has been frequently observed in various human diseases. Cui et al. demonstrated that TPM2 is often silenced due to abnormal DNA methylation, and its loss is linked to RhoA activation and cell proliferation^17^. Meng et al. reported that reduced TPM2 expression in atherosclerotic tissues accelerates apoptosis of vascular smooth muscle cells, leading to the release of multiple inflammatory factors and exacerbating intraplaque inflammation^18^. Shibata et al. found that TPM2 may play a critical role in inducing epithelial-mesenchymal transition (EMT) in the lens^19^. These findings collectively support our hypothesis that aberrant TPM2 expression might be associated with RCM in children.

Previous studies have highlighted the significant role of cardiac fibroblasts in RCM pathogenesis^20^. In vertebrate cardiac muscle, TPM interacts with troponin to centrally regulate calcium—dependent acto-myosin interactions, thereby controlling muscle contraction and relaxation^10^. Cardiomyocytes with abnormal sarcomeric structures—such as those involving troponin I—have been identified as key drivers of cardiac diastolic dysfunction^21^. Research has shown that *Drosophila* embryos lacking Tm2 exhibit reduced expression of sarcomeric proteins, while those expressing a gain-of-function Tm2 allele display defects in both cell fusion and elongation^22^.

Pathogenic variants in TPM2, which encodes a skeletal muscle-specific actin-binding protein essential for sarcomere function, significantly impair muscle development and function^5, 23^. A recent comprehensive review^24^ notes that infantile RCM can be associated with sarcomeric variants. Based on these findings, we hypothesize that TPM2 deficiency may lead to sarcomeric structural abnormalities in cardiomyocytes, ultimately contributing to cardiac diastolic dysfunction.

Our time-series analysis suggested that TPM2 expression is highest in naive cardiomyocytes, remains stable during maturation, and gradually decreases with cardiomyocyte aging—underscoring its critical role in cardiomyocyte development. Additionally, studies have reported that cardiomyocyte cell cycle activity contributes to postnatal cardiomyocyte proliferation, which is essential for postnatal growth and development^25^. Heart development is significantly affected by deletion of cyclin-D family members; mice with cyclin-D mutations exhibit ventricular malformations and hypoplastic ventricles^26^. In H9C2 cells, TPM2 knockdown resulted in S-phase cell cycle arrest, significantly reduced cyclin-D1 protein expression, and increased cardiomyocyte apoptosis. We therefore hypothesized that TPM2 may influence cardiomyocyte growth and development by regulating the cardiomyocyte cell cycle. Enhanced cardiomyocyte apoptosis leads to irreversible cardiomyocyte loss, reducing myocardial contractile units and worsening diastolic dysfunction. Given that Tm2 promotes F-actin assembly at the leading edge membrane^22^, the cellular mechanism of myotube elongation likely resembles that of migratory cells. We thus investigated the effect of TPM2 on myocardial cell migration, finding that TPM2 knockdown promoted myocardial fibroblast migration. Increased myocardial fibrosis can reduced ventricular compliance, impair cardiac contractile function, and potentially lead to heart failure^27^. Additionally, TPM2 knockdown significantly upregulated the expression of TGF-β1, α-SMA, and vimentin proteins, suggesting that TPM2 may regulate cardiomyocyte fibrosis via the TGF-β1 signaling pathway. Collectively, our findings identify a previously unrecognized role of TPM2: reduced TPM2 expression impairs cardiomyocyte proliferation, induces apoptosis, and promotes fibrotic progression.

This study has several limitations. First, our research focuses primarily on advanced pediatric RCM in patients eligible for transplantation. Future studies could explore whether more significant RCM-related differences emerge earlier in disease progression, though such investigations may be limited by sample availability. Second, the current study includes only participants of Asian descent; extending the research to diverse populations would enhance generalizability. Finally, the cardiomyocytes used to validate TPM2 function were derived from rats, which may introduce interspecies differences compared to human cardiomyocytes. Patient-derived induced pluripotent stem cell-derived cardiomyocytes (iPSC-CMs) are a powerful tool for cardiomyopathy research and would be particularly useful for delineating RCM pathogenic mechanisms, given the limited patient population.

In summary, this study provides a comprehensive characterization of the cellular transcriptional landscape in both healthy human hearts and those of pediatric RCM patients. Our analysis revealed significantly downregulated TPM2 expression in pediatric RCM heart tissues. We also demonstrated that TPM2 knockdown induces cell cycle arrest, triggers apoptosis, and accelerates fibrosis. These findings enhance our understanding of the transcriptional and molecular basis of pediatric RCM, offering insights that may inform future exploration of pathogenic pathways and potential therapeutic targets for this severe cardiac condition.

## Data Availability

The data that support the findings of this study are available from the corresponding author upon reasonable request.

## Non-standard Abbreviations and Acronyms

RCM: restrictive cardiomyopathy
TPM2: tropomyosin 2
TPMs: tropomyosins
scRNA-seq: single-cell RNA sequencing
snRNA-seq: single-nucleus RNA sequencing
EMT: epithelial-mesenchymal transition
HF: heart failure
LV: left ventricle
HT: heart transplantation
IRBs: Institutional Review Boards
GO: Gene Ontology
BP: biological processes
CC: cellular components
LC-MS/MS: Liquid chromatography-tandem mass spectrometry
TMT: tandem mass tags
NC: negative control
siRNA: small interfering RNA
FITC: fluorescein isothiocyanate
PI: propidium iodide
DAPI: 4’,6-diamidino-2-phenylindole
FBS: fetal bovine serum
DMEM: Dulbecco’s Modified Eagle Medium
RT-qPCR: Quantitative real-time PCR
WB: Western blotting
BSA: bovine serum albumin
PPI: protein-protein interaction
iPSC-CMs: induced pluripotent stem cell-derived cardiomyocytes

## Author Contributions

Jie Liu and Zubo Wu conceptualized, designed, performed, and anlyzed the experiments and wrote the article; Cong Zao performed and analyzed experiments; Qing Guo performed experiments; Nianguo Dong and Peng Zhu designed experiments and provided critical reagents; Jiawei Shi, Lin Wang; and Hua Peng designed experiments, provided critical input and revised the article.

## Acknowledgments

We are grateful to Dr. Lingqiang Zhu from the Department of Pathophysiology, School of Basic Medicine, Tongji Medical College, Huazhong University of Science and Technology, for his valuable advice on experimental design and technical assistance. We acknowledge the support of the Ethics Committee of Union Hospital for approving this study (Approval No. KY2017-323), and express our thanks to all participating patients and their families for donating clinical samples.

## Sources of Funding

Natural Science Foundation of Hubei Province, China(2023AFB494); National Key R&D Program of China(2023YFC2706200); Wuhan Knowledge Innovation Special Project (2023020201010160); key research and development program by Hubei province of China (2022BCA043); Natural Science Foundation of Hubei Province of China (2020CFB813); Supported by Natural Science Foundation of Hubei Province of China (2020CFB764)

## Disclosures

None

## Notes

### Competing Interest Statement

The authors have declared no competing interest.

### Author Declarations

Ethics Committee of Union Hospital(Approval No. KY2017-323)

